# Using portable air purifiers to reduce airborne transmission of infectious respiratory viruses – a computational fluid dynamics study

**DOI:** 10.1101/2021.11.01.21265775

**Authors:** Liwei Guo, Ryo Torii, Ruth Epstein, John Rubin, Jonathan P. Reid, Haoran Li, Andrea Ducci, Ramanarayanan Balachandran, Manish K. Tiwari, Yiannis Ventikos, Laurence B. Lovat

**Author notes:** Corresponding author. E-mail address (Liwei Guo), (Ryo Torii).

## Abstract

Aerosols and droplets generated from expiratory events play a critical role in the transmission of infectious respiratory viruses. Increasingly robust evidence has suggested the crucial role of fine aerosols in airborne transmission of respiratory diseases, which is now widely regarded as an important transmission path of COVID-19. In this report, we used CFD modelling to investigate the efficiency of using portable air purifiers containing HEPA filters to reduce airborne aerosols in hospitals and serve as a potential retrofit mitigation strategy. We used a consulting room to set up our simulations because currently the clearance time between consultations is the controlling factor that limits the patient turnover rate. The results suggest the inlet/suction of the air purifier unit should be lifted above the floor to achieve better clearance efficiency, with up to 40% improvement possible. If multiple air purifiers are used, the combined efficiency can increase to 62%. This work provides practical guidance on a mitigation strategy that can be easily implemented in an expedient, cost-effective and rapid manner, and paves the way for developing more science-informed strategies to mitigate the airborne transmission of respiratory infections in hospitals.

## 1 Introduction

Aerosol particles and droplets generated from expiratory events, such as breathing, talking, singing, coughing and sneezing [1,2], play a critical role in the transmission of infectious respiratory diseases, such as SARS (Severe Acute Respiratory Syndrome), MERS (Middle East Respiratory Syndrome) and COVID-19 (Coronavirus Disease 2019). To understand how far and fast droplets/aerosols can travel in indoor spaces is essential for developing effective mitigation strategies for infection control, especially in high-risk environments, such as hospitals. Normally, large droplets settle to surfaces over short distances (< 2 m); intermediate droplets deviate from the exhaled jet and evaporate to an equilibrium size; and small droplets travel with the exhaled air eventually also drying to a composition and size governed by the ambient conditions (relative humidity) [3]. Droplets in the last category, also defined as fine aerosols (aerodynamic diameter ≤ 5 μm), remain airborne for much longer and can spread much further than larger droplets [4,5].

There is evidence that suggests fine aerosols play a crucial role in the airborne transmission of respiratory diseases [6], and aerosol transmission is now widely regarded as an important transmission path of COVID-19 [2]. Ventilation, particle settling and air disinfection have been suggested as viable mitigation mechanisms [7]. Since aerosol dispersion strongly follows convective air flows [4], mitigation measures involving ventilation are shown to provide more efficient solutions, such as higher air exchange rates [2,8] and introducing flow-directing geometries [9].

However, substantial redesign of the entire Heating, Ventilation and Air Conditioning (HVAC) systems might be expensive, whereas portable air purifiers containing high-efficiency particulate absorbing (HEPA) filters, which filter more than 99% of particles larger than 200 nm [10], have the potential to provide a cost-effective, retrofit solution with several advantages, such as flexible positioning, easy maintenance and user-friendly control interfaces.

Although there is evidence to support using portable air purifiers to disinfect polluted air [11], there is little data to demonstrate the most effective usage. This raises a few key questions – Where should we put them in a room? For a given room and existing HVAC setting, do we need to use the maximal flow rates in the purifiers? Higher flow rates can filter more particles, but the stronger outflow jet may disturb the airflow and cause unwanted mixing, not to mention the increased noise level. Here we report a summary of our computational study on the performance of portable air purifiers based on Computational Fluid Dynamics (CFD) simulations. CFD has been extensively applied in studies of aerosol generating activities [12] and ventilation in hospitals [13]. We used the geometry and setup of a consulting room as a starting point to set up the simulations because currently the clearance time between consultations is the controlling factor that limits the patient turnover rate. In this report, we present our CFD modelling results of aerosol dispersion in hospitals and the reduction after introducing portable air purifiers, which offers characterisation of the performance and provides practical guidance to maximise efficiency.

## 2 Materials and methods

We created a model representing a typical consulting room in a UK hospital, with one doctor and one patient in sitting postures. The dimensions of the room are shown in Figure 1a (length: 5.6 m, width: 2.5 m, height: 2.7 m). We generated a computational mesh using non-uniform structured hexahedral elements, with local refinement at surfaces. A detailed mesh convergence study was conducted on four different mesh sizes, and the one used in this report has around 800,000 elements. We performed 3D transient simulations, using a time step of 0.01 s, over the time span of 180 s. Both the continuous phase (air flow) and the discrete phase (aerosols) were taken into account in the simulations. The continuous medium was air at atmospheric conditions (density *ρ* = 1.16 kg/m^3^, reference pressure *p* = 101,325 Pa). The air flow and temperature field were simulated by solving 3D incompressible Navier-Stokes and heat transfer equations using the finite volume method with SIMPLEC pressure correction [14]. The standard *k*-*ε* model was used to account for the effects of turbulence [15]. Gravity and buoyancy forces were fully accounted for. A variety of schemes, of different degrees of spatial and temporal accuracy, were used to ascertain the sensitivity of these specific results to such numerical factors. The discrete phase of aerosols was tracked in a Lagrangian manner, with representative populations of around 200,000 particles being tracked. The sensitivity of the results and conclusions on the number of representative particles tracked was also confirmed. The particle drag force is a function of the local Reynolds number. The material of aerosols was assumed to be water at atmospheric conditions, and they were set to adhere to the wall upon contact. We used a fixed diameter of 1 μm (single size) to represent fine aerosols and evaporation was not considered based on the experimental evidence on aerosol particle diameter equilibration times [4]. In effect, we assumed the size distribution of such small aerosol was already equilibrated to the environmental conditions at the first step of the simulation and only heat but no mass transfer between the aerosols and the air flow occurred. The difference in temperature between the human models, and therefore the exhaled air and aerosols, and the ambient temperature created buoyant (initially rising) plumes that affected the particle distributions and were fully accounted for. We also used a stochastic model for turbulent dispersion of particles [16]. In the simulations, viscous drag, turbulent flow dispersion and buoyancy are the main forces that determine the trajectories of aerosols. Although the void fraction of particles is small, we chose to employ two-way (i.e. full) momentum coupling between particles and fluid, to enhance the accuracy of the results and to prepare for further developments regarding the physics included, like evaporation of droplets.

**Figure 1:**
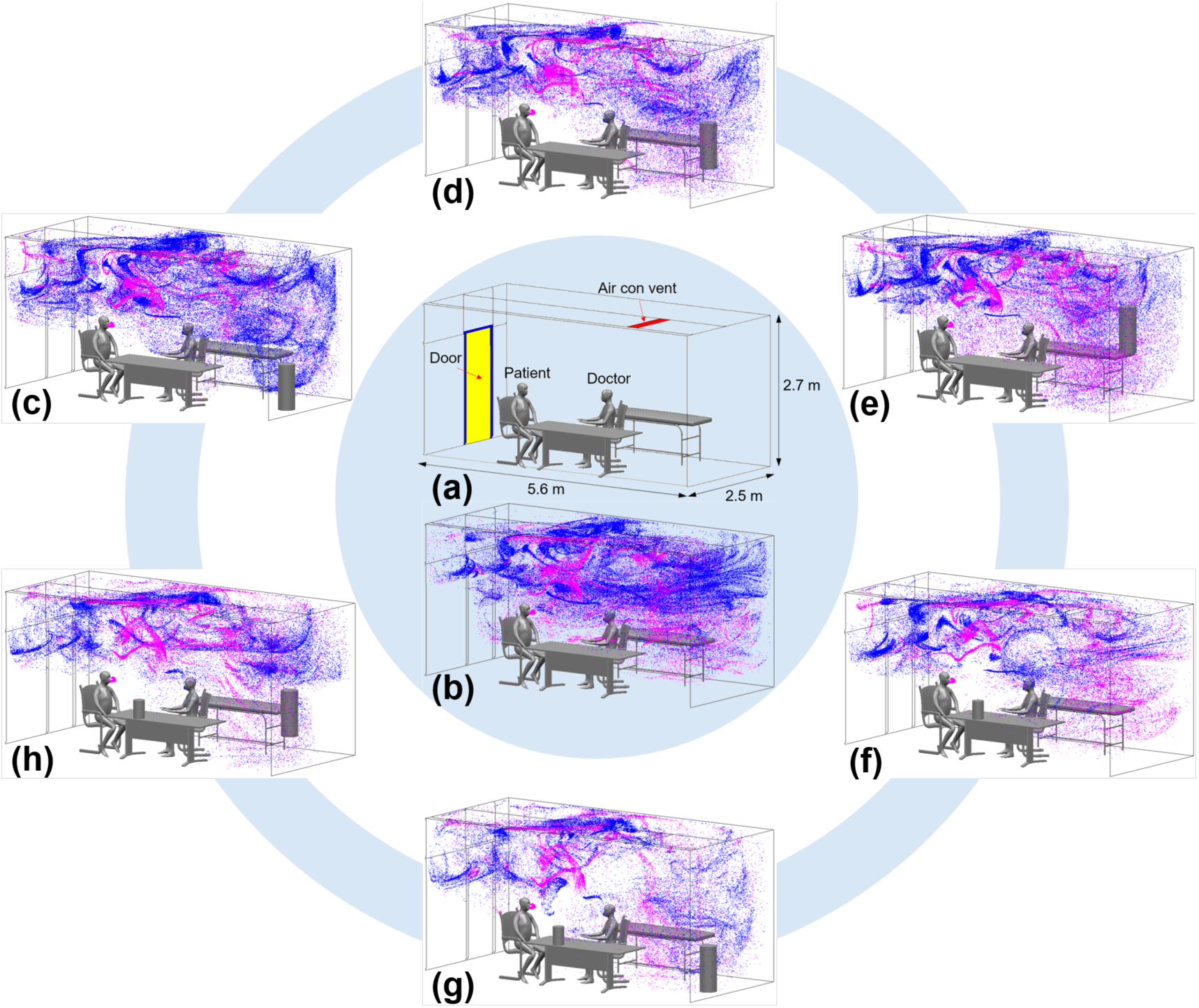
Aerosol dispersion in different scenarios for a hospital consulting room. Blue aerosols are from the doctor (sitting on the right) and magenta aerosols are from the patient (sitting on the left). (a) The consulting room geometry and dimensions. (b) The baseline simulation, where only the air conditioning is working. (c) In addition to the air conditioning, AP-I is placed on the floor in the back empty space of the room. (d) At the same horizontal position, AP-I is lifted 0.5 m above the floor. (e) AP-I is further lifted 1 m above the floor. (f) The air conditioning is working at the same condition, but AP-II is placed on the desk instead of AP-I in the back of the room. (g) A combined solution – both AP-I on the floor and AP-II on the desk. (h) AP-I is lifted 0.5 m above the floor whereas AP-II stays at the same position on the desk.

Previous research suggested that normal breathing and talking are responsible for a portion of the virus-laden aerosols [17]. Therefore, we prescribed time-varying velocity boundary conditions at the mouth of the human models. More specifically, the patient was assumed to be breathing in a normal way (12 breaths per minute), *v*_*p*_ = 2.8 × sin(1.26*t*); and the doctor was assumed to be talking (15 breaths per minute), *v*_*d*_ = 2.8 × sin(1.57*t*), where *t* is time in seconds [18]. The door was closed (yellow area in Figure 1a), but air was allowed to leak from small gaps around the door (blue strips in Figure 1a), where a fixed (atmospheric) pressure boundary condition was applied. There was only one vent of the air conditioner on the ceiling, with a constant vertically downward velocity of 0.16 m/s, which is equivalent to an air change rate of 3/hour and is the normal working condition for clinical consulting rooms [13]. No-slip boundary conditions were applied at the walls, floor, ceiling and human models. Rectangular openings were cut around the mouth of human models to inject aerosols with the exhaled air at a constant rate depending on the local mesh. The room temperature was 23 °C and the human body temperature was set at 37 °C. Two types of air purifiers containing HEPA filters were simulated – both having a cylindrical shape, sucking air in from the lateral surface of the lower half and supplying fresh filtered air from the top; in the simulations we assumed 100% filtration efficiency so there were no aerosols injected from the top. The large one (Air Purifier I, AP-I) is 80 cm high (diameter 30 cm), with a medium air flow rate of 300 m^3^/h; the small one (Air Purifier II, AP-II) is 32 cm high (diameter 20 cm) and has a medium air flow rate of 65 m^3^/h. We used CFD-ACE+ (ESI Group, Paris, France) for all of the simulations; each simulation ran in parallel on 16 cores.

## 3 Results and discussion

Figure 1 shows the aerosol dispersion in the consulting room at *t* = 180 s; the baseline simulation is shown in the centre and different scenarios of using portable air purifiers are arranged in a circular manner. We cut off the simulations at 180 s because it can be seen from the plots in Figure 2 that the aerosol particle number reaches a plateau around that time, which therefore can be assumed to represent a steady state.

**Figure 2:**
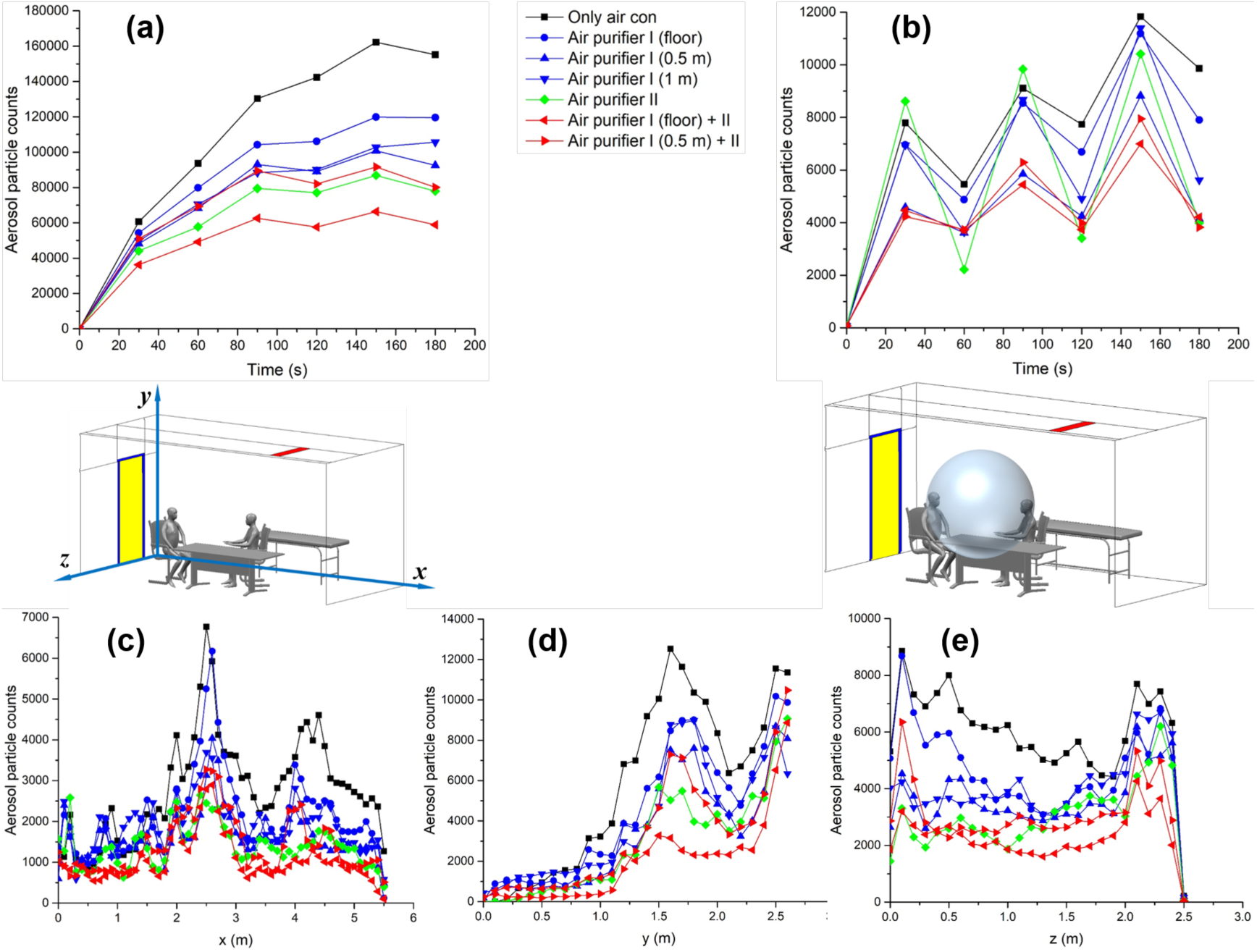
Plots of temporal and spatial distributions of aerosol particle counts. (a) Particle counts in the entire room vs. time. (b) Particle counts within a local spherical region between the two human models. (c) Particle count distribution in the *x*-direction at *t* = 180 s. (d) Particle counts in the vertical *y*-direction at *t* = 180 s. (e) Particle counts in the *z*-direction at *t* = 180 s.

Figure 2 shows the spatial and temporal distributions of aerosol numbers. It should be noted that in the simulations we used normalised mass for the injected aerosols, which means a fixed number of aerosols at the beginning (*t* = 0) for all the simulations and we focus on the relative differences between various scenarios, not the absolute numbers. The number of aerosols was counted both globally in the entire room and in a local spherical region of diameter 1.6 m with its centre at the same vertical level as the mouth of the human models, to represent the region of room from which aerosols might be inhaled by the patient or doctor. Because the breathing velocities of human models were assumed to follow sinusoidal curves, we observe clear fluctuations in the local spherical region. The spatial distributions of the particles were also analysed at the end of the simulations (*t* = 180 s), where particle counts were binned in slabs of 0.1 m in the *x, y* and *z* directions, respectively.

The baseline simulation (Figure 1b) shows a markedly dense aerosol concentration in the upper part of the room, mainly due to the thermal plume effect, which can also be seen in the plots of Figure 2d, where particle counts in the vertical *y*-direction increase dramatically above the level of the human model mouth (approximately 1.2 m). We also see more blue aerosol particles (from the doctor sitting on the right) than the magenta ones (from the patient sitting on the left) because the doctor was assumed to be talking so exhaled more aerosols, which was implemented as higher aerosol concentration from the mouth in the simulations. Then, we added portable air purifiers into the simulations. First, we simulated Air Purifier I (AP-I). Normally, this type of air purifier is placed on the floor; however, from the baseline simulation we can see more aerosols floating above the breathing level thus we compared different vertical positions – on the floor (Figure 1c), 0.5 m above the floor (Figure 1d) and 1 m above the floor (Figure 1e). Since the air purifier AP-I is 0.8 m tall, it is not practical to place it any higher. Reduction factors were computed to quantitatively compare different scenarios, which are calculated as the ratio of the aerosol particle counts at *t* = 180 s between the baseline simulation and the mitigation scenarios. The results show that using AP-I can effectively reduce the relative aerosol numbers achieved at steady state in the entire room, to 0.77 (on the floor), 0.60 (0.5 m above the floor) and 0.68 (1 m above the floor), respectively (Figure 2a); locally, a reduction factor of 0.80, 0.42 and 0.57 were obtained for the three heights (Figure 2b). The results are naturally generalisable and suggest that portable air purifiers can be used as a retrofit mitigation strategy to reduce airborne aerosols. From a practical point of view, elevating air purifiers above the floor is more effective in clearing aerosols than placing them directly on the floor.

Next, the smaller air purifier II (AP-II) was placed on the desk (Figure 1f) and a reduction factor of 0.50 was observed globally and 0.40 locally at steady state. Then, we combined the two air purifiers, with the large one in the slightly far-field empty space and the small one closer to the sitting area (Figure 1g, h), which can further mitigate a reduction factor of around 0.38 (AP-I on the floor) and 0.52 (AP-I 0.5 m above the floor) in the entire room, and 0.43 and 0.39 in the local spherical region. The results suggest that using multiple air purifiers can be more effective, but their relative positions are important. In this case, the large purifier works better on the floor together with the small one on the desk. This is because there is less mixing of the jets from different air purifiers to disturb each other’s suction zone (Figure 1g, h), which turned out to be the most effective mitigation solution simulated in this report. Compared with the baseline simulation, where only the air conditioning is working, using air purifiers is equivalent to increasing the air change rate. Depending on the energy requirement of the central HVAC system, using portable air purifiers may be more energy-efficient.

This short report summarises our CFD modelling work so far; however, more research still needs to be done, such as validation with experiments and generalisation to other settings. From the infection control perspective, statistical models can be introduced to model uncertainty and therefore predict the risk of exposure [19]; and quantitative models to link aerosol concentration and probability of infection are also worth exploring [9].

## 4 Conclusions

We used CFD modelling to investigate the efficiency of using portable air purifiers to clear fine aerosols in hospitals. The results suggest that air purifiers can effectively reduce airborne aerosols. More specifically, the inlet of suction should be lifted above the floor to achieve higher reduction in steady-state aerosol concentration (40%); and using multiple air purifiers can further increase the efficiency up to 62%. This work provides practical guidance on a mitigation strategy that can be easily implemented, and paves the way for developing more science-informed strategies to mitigate the airborne transmission of respiratory infections in hospitals, especially for high-risk and aerosol-generating procedures, such as endoscopy, dentistry and speech therapy.

## Data Availability

This article has no additional data.

## Data accessibility

This article has no additional data.

## Authors’ contributions

LG: study conceptualisation, study design, data collection, data analysis, data interpretation, manuscript preparation, manuscript revision.

RT: study conceptualisation, study design, data analysis, data interpretation, manuscript revision, supervisory oversight.

RE, JR, JPR: study conceptualisation, data interpretation, manuscript revision.

HL: study design, data collection.

AD, RB: study conceptualisation, study design, data interpretation, manuscript revision.

MKT, YV, LBL: study conceptualisation, study design, data interpretation, manuscript revision, supervisory oversight.

## Acknowledgements

LG and YV would like to acknowledge the financial and technical support provided by the ESI Group in association with the use of its CFD-ACE+ software package throughout this research. LBL is supported by the National Institute for Health Research University College London Hospitals Biomedical Research Centre and the Wellcome/EPSRC Centre for Interventional and Surgical Sciences (WEISS) at UCL; [203145Z/16/Z]. The team wish to thank Dr Gee-Yen Shin and Dr Catherine Houlihan, Consultant Virologists, and Diana Kootstra, Senior Construction Programme Manager at University College London Hospitals NHS Foundation Trust and Matthew Day, Regional Director at DSSR Limited for their support. Support from UCL Mechanical Engineering is also gratefully acknowledged.

